# Radiation-specific Automated Dosimetric dental, Mandible, and maxilla Annotation for Predicting Periodontal Problems (RADMAP): A semi-automated tool for dosimetric oral risk communication and osteoradionecrosis assessment

**DOI:** 10.1101/2025.11.23.25340237

**Authors:** Lisanne V van Dijk, Laia Humbert-Vidan, Erin E Watson, Ruth A Aponte Wesson, Luisa E Jacomina, Renjie He, Mohamed A Naser, Dong Joo Rhee, He C Wang, Stephen Y Lai, Muhammad Walji, Max JH Witjes, Matthew S. Katz, Andrew Hope, Mark S Chambers, Clifton D Fuller, Amy C Moreno

**Affiliations:** Department of Radiation Oncology, University Medical Center Groningen, Groningen, NL; Department of Radiation Oncology, The University of Texas MD Anderson Cancer Center, Houston, USA; Department of Dental Oncology/Faculty of Dentistry, Princess Margaret Cancer Centre; University Health Network; University of Toronto, Toronto, CA; Department of Head and Neck Surgery, The University of Texas MD Anderson Cancer Center, Houston, USA; Department of Radiation Physics, The University of Texas MD Anderson Cancer Center, Houston, USA; Department of Clinical and Health Informatics, UTHealth Houston McWilliams School of Biomedical Informatics; Oral & Maxillofacial Surgery, University Medical Center Groningen, Groningen, NL; Department of Radiation Oncology, Tufts Medical Center, Boston, MA; Department of Radiation Oncology, University of Toronto, Toronto, Ontario, Canada

## Abstract

**Purpose/Objective(s):** Osteoradionecrosis of the jaw (ORN) is a severe complication of head and neck cancer (HNC) radiotherapy (RT), significantly impacting patient quality of life. Current dose assessment relies on whole-mandible dosimetry, limiting personalized, tooth-specific region risk evaluation. The lack of standardized methods to document and analyze dose distributions to tooth-bearing regions further hampers dental and maxillofacial decision-making. This study develops and evaluates RADMAP, a (semi-)automated tool for segmenting tooth-based jaw regions, enabling patient- and tooth-specific radiation dose mapping to improve dental dose reporting and ORN risk assessment.

**Methods and Materials:** A total of 736 tooth locations from 23 HNC patients treated with definitive RT were analyzed, including 11 who developed ORN. The RADMAP tool applies an angular ray-based algorithm to automatically segment the mandible and maxilla into 32 tooth-specific jaw regions, with optional manual refinement (semi-automated), outputting radiation dose mapping in both tabular and odontogram formats. Mean dose values from manually contoured tooth roots were compared with RADMAP-segmented alveolar regions (fully and semi-automated). Interobserver agreement among six users was evaluated. Whole-mandible, manual tooth, and tooth-based jaw segment (alveolar and basal) doses were compared between regions with and without ORN development.

**Results:** Mean dose correlation between RADMAP-segmented alveolar jaw regions and manually segmented tooth roots was high for both fully and semi-automated approaches (R² = 0.98, p<0.0001), demonstrating accurate dose estimation. Interobserver agreement showed 95% limits of ±2.9 Gy for mandibular alveolar segments, confirming reproducibility. Tooth-based jaw segments showed a significant differentiation in radiation dose for the ORN-positive versus ORN-negative sites (difference in mean dose (DIM): 12.4 ±3.7 Gy, p=0.0008), which was not seen when considering the whole-mandible dose (DIM=6.2 ±4.3 Gy, p=0.17), demonstrating RADMAP’s promise for improved ORN risk assessment.

**Conclusion:** RADMAP enables accurate, tooth-specific dose mapping of the mandible and maxilla, and potential improved ORN risk differentiation beyond whole-mandible dosimetry. Tooth-based dose reporting can personalize treatment, enhance multidisciplinary communication, and support prevention of radiation-related orodental sequelae.

## INTRODUCTION

Radiation therapy (RT) plays a pivotal role in head and neck cancer (HNC) management and is associated with several orodental toxicities, including xerostomia, periodontal disease (POD), post-RT caries (PRC), and osteoradionecrosis of the jaw (ORN)^1^. ORN, in particular, is one of the most severe toxicities that can develop, significantly impacting patient quality of life through orofacial functional impairment, pain, increased burden of chronic dysphagia, fracture, and costly surgical interventions^2^.

Measures to reduce the ∼1 in 10 incidence of ORN in HNC survivors treated in the modern era are a high priority, as emphasized by recent ISOO-MASCC-ASCO guidelines^3^. These include pre-therapy dental evaluations and prophylactic extractions when indicated, aiming to minimize post-RT interventions, which are strongly associated with ORN development^3,4^. This clinical decision-making process often considers the existing dental condition, such as the presence of infection foci, in conjunction with the anticipated high radiation doses to specific tooth regions (e.g., often an arbitrary 30 Gy threshold)^5^. However, there is a clinical absence of effective methods for documenting and analyzing dosimetric distributions to tooth-bearing regions which impedes dentists and maxillofacial surgeons from making data-driven decisions for personalized, toxicity-mitigating dental care in the pre- and post-RT settings.

Current studies examining relationships between dental procedures, radiation dose, and RT-related sequelae largely rely on dichotomized features (e.g., “yes” or “no” for pre-RT dental extractions) and whole organ (i.e., mandible) dosimetry to identify significant dose-volume correlates of ORN. A previously developed normal tissue complication probability (NTCP) prediction model based on a large-scale observational cohort (n=1259) showed that limiting 30% of the mandible to receiving 35 Gy (in patients with pre-radiotherapy extractions) or 42 Gy (in patients without pre-radiotherapy extractions) or less was associated with a <5% ORN risk^6^. Additionally, other studies identified similar dosimetric parameters of the mandible, including V58Gy <25% and mean dose <37Gy for use during RT planning^3,7,8^. While current ORN prediction models are useful for guiding RT decision-making and surveillance strategies, they provide limited information for targeted dental care due to their lack of tooth-specific dose relationships, emphasizing the necessity for comprehensive assessment of tooth-region specific dose data to better inform dental care policies and investigate ORN development at a localized level.

Auto-segmentation of teeth on dental CT or cone beam CT images has been a recent development in dental research^9^. Traditional segmentation methods such as atlas-based segmentation^10,11^ or deep learning^12,13^ have shown excellent performance at auto-delineating the mandible structure. However, a translational gap exists in applying tooth-autosegmentation approaches to planning CT scans linked to RT dose distribution plans, which are necessary to extract tooth-specific dose parameters. A limitation for current segmentation approaches (e.g., deep learning) is that they cannot segment volumes-of-interest that are absent^14^, as HNC patients often have had teeth extracted and/or are partly or fully edentulous. Hence, there is a need for an efficient teeth region segmentation method that is agnostic to the presence of teeth.

The primary objective of this study was to develop a vendor-independent clinical tool for semi-automated segmentation of tooth-bearing regions of the jaw, maxilla included, with generation of automated reports translating dose data onto a ubiquitous dental assessment tool (odontogram) with associated tabular data. We hypothesized that autosegmentation of tooth-bearing subregions of the jaw using an angular ray-based algorithm could be reliable and universally applied to all HNC patient RT planning CT scans, and that these subregions (i.e., alveolar bone versus jaw segments) could be useful for analysis of localized tooth-specific dose relationships with ORN development. Herein, we report 1) the technical development and output of the RADiation dose MAPping tool [RADMAP], 2) the interrater reliability results, and 3) the tool application in localized radiation dose analysis to identify ORN.

## METHODS

### Patient cohort

After Institutional Review Board approval [RCR03-0800, 2024-002], retrospective treatment and imaging data were collected from 23 patients with newly diagnosed head and neck squamous cell carcinoma (HNSCC) who received definitive or postoperative RT from 2006 to 2020 at a single institution. Patients were selected based on having most of their natural dentition, a planning CT scan without substantial metal artifacts, and with approximately half having developed ORN (see section ORN dosimetric analysis). Patient characteristics are summarized in Table 1. Overall, most patients were male (91%), 44% were former smokers with a median of 40 pack years (range: 3-54), and 74% reported alcohol use (current or former). Tumors were primarily located in the tongue base (57%) or tonsil (35%), surgery was performed on 3 (13%) patients, and systemic therapy was delivered in 87% of patients. The RT prescription dose ranged from 65–70 Gy delivered in 30-35 fractions, 48% of the cohort received “split-field” intensity modulated RT (IMRT) or volumetric arc therapy (VMAT), and 74% were treated with a customized mouth-opening, tongue-deviating dental stent in place^15^. Split-field IMRT/VMAT plans generally treated the mid-low neck with a matched larynx block field to 50 Gy with a full midline block to shield the spinal cord after 40 Gy.^16^ With regards to pre-therapy dental evaluations, the majority had either good or fair oral hygiene (87%) and 22% were recommended to have pre-RT extractions. This study was approved by The University of Texas MD Anderson Cancer Center Institutional Review Board under the following data collection and analysis protocols: PA14-0947, RCR03-0800, and 2024-0002).

**Table 1.** Cohort and Treatment Characteristics.

| Characteristic | Overall<br>(N=23) |
| --- | --- |
| Age (Years) |  |
| Mean (SD) | 56.6 (9.48) |
| Median [Min, Max] | 58.0 [35.0, 73.0] |
| Sex |  |
| Male | 21 (91.3%) |
| Female | 2 (8.7%) |
| Smoking History |  |
| Never smoker | 13 (56.5%) |
| Former | 10 (43.5%) |
| Smokeless Tobacco Use |  |
| Yes | 2 (8.7%) |
| No | 18 (78.3%) |
| Unknown | 3 (13.0%) |
| Alcohol Use |  |
| Yes | 17 (73.9%) |
| No | 6 (26.1%) |
| Primary Tumor Site |  |
| Base of tongue | 13 (56.5%) |
| Tonsil | 8 (34.8%) |
| Oral tongue | 1 (4.3%) |
| Neck (Unknown Primary) | 1 (4.3%) |
| Tumor Laterality |  |
| Right | 14 (60.9%) |
| Left | 9 (39.1%) |
| T Stage (7th) |  |
| T0 | 1 (4.3%) |
| T1 | 2 (8.7%) |
| T2 | 9 (39.1%) |
| T3 | 6 (26.1%) |
| T4 | 5 (21.7%) |
| N Stage (7th) |  |
| N1 | 2 (8.7%) |
| N2a | 2 (8.7%) |
| N2b | 13 (56.5%) |
| N2c | 6 (26.1%) |
| RT Total Dose (cGy) |  |
| Mean (SD) | 6900 (174) |
| Median [Min, Max] | 7000 [6510, 7000] |
| RT Total Fractions |  |
| Median [Min, Max] | 33 [30, 35] |
| RT Modality |  |
| IMRT/VMAT - Split field | 11 (47.8%) |
| IMRT/VMAT - Full field | 9 (39.1%) |
| IMPT | 3 (13.0%) |
| Dental Stent during RT |  |
| Yes | 17 (73.9%) |
| No | 6 (26.1%) |
| Surgery |  |
| Yes | 3 (13.0%) |
| No | 20 (87.0%) |
| Systemic Therapy |  |
| Concurrent only | 11 (47.8%) |
| Induction-Concurrent | 7 (30.4%) |
| Induction only | 2 (8.7%) |
| None | 3 (13.0%) |
| Pre-RT Extractions Recommended |  |
| Yes | 5 (21.7%) |
| No | 18 (78.3%) |
| Pre-RT Oral Hygiene |  |
| Good | 15 (65.2%) |
| Fair | 5 (21.7%) |
| Poor | 1 (4.3%) |
| Not reported | 2 (8.7%) |

### Manual orodental structure contouring

For each patient, individual teeth were manually delineated and labeled using the universal tooth numbering system (1 to 32) on the non-contrast radiotherapy planning CT (Supplementary data S1)^17^. The entire mandible (including condyles) and maxillae were manually contoured as separate organs-at-risk (OARs) using consensus-based delineation guidelines.^18,19^ All manual contours were reviewed and approved by ACM.

### RADMAP tool design

#### Multidisciplinary expert Delphi study and tool functionality

Our recent international Delphi study (n = 69) informed the RADMAP tool’s visual design^20^, showing strong expert support for an open-mouth odontogram with heat-map visualization and inclusion of both maximum (d_max_) and mean (d_mean_) radiation dose parameters (Supplement S1 for full details). The tool’s main objectives were to automatically overlay 3D dose distributions onto an odontogram, calculate and display d_mean_ for individual tooth-bearing regions in tabular form, and enable manual adjustment of tooth-region borders for improved dosimetric precision.

#### RADMAP design process

An overview of the RADMAP tool and its output is shown in Figure 1. The tool, developed in MATLAB R2023a (MathWorks Inc., Natick, MA), launches via a terminal prompt that requests the input directory containing the patient DICOM data and the desired output directory. All patients with a complete DICOM set (CT, RTDOSE, and RTSTRUCT files) are listed for selection. Upon selection, the corresponding CT, RTDOSE, and RTSTRUCT files—containing at least the mandible and optionally the maxilla—are loaded, and all data are matched to the CT.

**Figure 1.**
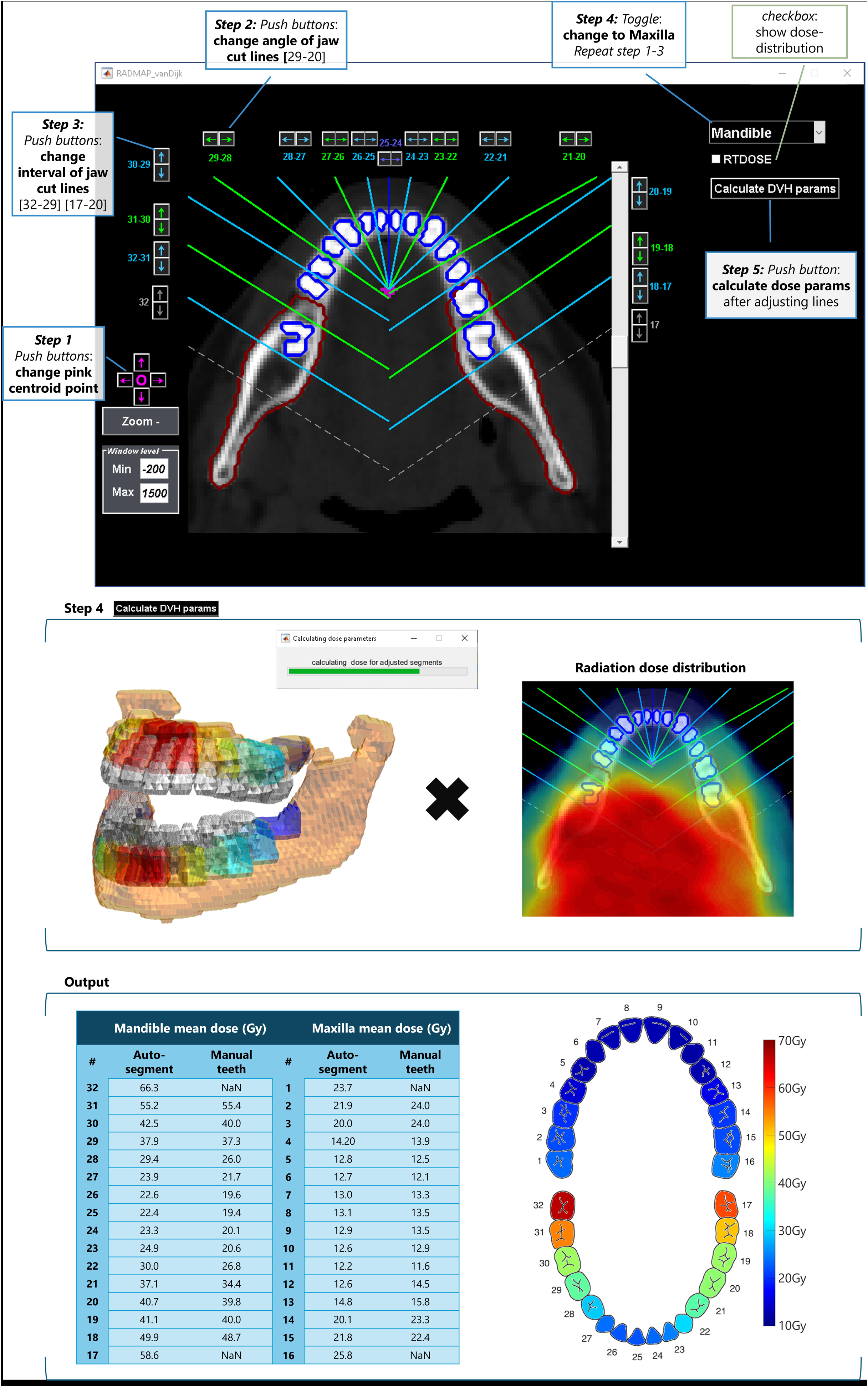
A) The RADMAP user interface showing manual adjustment options for (1) the centroid, (2) individual delimiters, (3) delimiter angles, and (4) switching views between mandible and maxilla, prior to (5) calculation of dental dosimetric parameters. B) a 3-dimensional representation of teeth-based jaw segments and the radiation dose distribution overlayed on a CT image. C) the output tabular display of d_mean_ per teeth section and the radiation odontogram.

The jaw centroid and spatial dimensions are derived from the mandible and maxilla (if available) contours in the CT images. The CT is then centered and cropped to the jaw boundaries. A *CenterPoint* (pink dot, Figure 1A) is defined at one-third of the distance between the centroid and the anterior jaw limit. Bilateral jaw delimiters (dashed gray lines) are automatically placed at the mandibular angle—the point of maximum gradient change along the mandible projection—marking the posterior molar boundary. For the mandible, the search range extends from the CenterPoint to 1.5 cm anterior to the posterior limit, while for the maxilla, delimiters are set at the posterior contour margins.

Incisor delimiters were defined by first placing a midline delimiter (dark blue line in Figure 1) through the CenterPoint, followed by additional delimiters computed using the tangent function at angles of [−49°, - 29°, −13°, 0°, 13°, 29°, 49°] for the mandible and [−58°, −40°, −20°, 0°, 20°, 40°, 58°] for the maxilla (light blue and green lines in Figure 1).

To determine the optimal angulation of delimiters separating premolars and molars, a summed 2D projection was generated for each jaw half (e.g., right side), spanning from the last front-tooth angle (49°) to the jaw end. For the mandible, only the bone visible in ≥3 slices was included. A linear regression fitted through the resulting coordinate cloud provided a slope, which was converted via inverse tangent to the (pre-)molar delimiter angle. Molar delimiters were positioned using this angulation with interval spacings of [2%, 27%, 50%, 75%, 100%] for the CenterPoint–BendPosition distance for the mandible, and [12%, 23%, 54%, 80%, 100%] for the maxilla.

To auto-delineate jaw segment volumes, the volume of jawbone between tooth-based delimiters was converted to a binary mask with a label corresponding to the tooth index. Total jaw segments were extracted, as well as an autosegmentation of the alveolar (i.e., root-bearing) versus basal regions of each jaw segment (Figure 1B). The d_mean_ for each jaw segment was calculated using linked RTDOSE files. These dental dosimetric parameters were then exported as a tabular output and an odontogram overlayed with a heatmap of d_mean_ per individual tooth (Figure 1C). Additional output files included data on coordinates, other dose-volume (DVH) parameters, and 3D figures (see Supplement Figure S2).

Manual functions within the RADMAP tool enable fine-tuning of delimiter placement, thereby improving its applicability across diverse cohort settings. These steps are illustrated in steps 1–5 of Figure 1A.

#### RADMAP evaluation

In total, six observers (2 radiation oncologists [ACM, LEJ], 2 dental specialists [RAAW, EW], a medical physicist [LHV], and a technical physician [LVD]) loaded the same 23 cases into the RADMAP tool for independent review, manual adjustments, and exporting of RADMAP data.

### Manual tooth versus RADMAP jaw segment analysis

The manually contoured tooth roots were defined as the portion of the manually segmented tooth intersecting the mandible or maxilla, while the alveolar segment represented a 1.5 cm vertical expansion from the most superficial tooth-bearing bone—bounded laterally by delimiters (Figure 1B). This alveolar bone height estimation was informed by expert CT review and published reference ranges^21–23^, and was essential to enable alveolar jaw segmentation in new patients without requiring individual tooth segmentations.

To evaluate whether the RADMAP tool accurately estimates radiation dose to tooth roots, mean doses to the root and alveolar jaw segment were compared for both fully automated and semi-automated (i.e., manually adjusted) segmentations, averaged across observers. Volume and geometric overlap comparisons were performed, and linear correlation analyses were used to assess the relationship between mean dose estimates from automated and manual delineations. Agreement between methods was evaluated using Bland–Altman analysis, reporting systematic bias, standard deviation (SD), and 95% limits of agreement (LOA). Statistical significance was set at p < 0.05.

### Inter-observer analysis

To assess the consistency in jaw segment estimation by different users of the tool, the d_mean_ of the alveolar jaw segments of all tooth indexes were compared to the average across all observers. Bland-Altman analyses were conducted to determine the 95% LOA. These analyses provided a measure of inter-observer consistency in segment delineation.

### Case application: ORN dosimetric analysis

Building on our previous studies linking full mandibular DVH parameters to ORN risk^6,7^, a spatial analysis was performed to evaluate ORN development relative to clinically relevant jaw-segment doses. The d_mean_ and d_max_ were extracted for the entire mandible, manually segmented tooth roots, and fully auto-delineated jaw, alveolar, and basal bone segments (Supplement Figure S3). In 11 patients who developed ORN after radiotherapy, the ORN location was identified on surveillance CT imaging and recorded according to the involved tooth indices. Dosimetric parameters of ORN-affected regions were compared with unaffected teeth *within* the same patient and with all teeth from patients without ORN using difference in mean (DIM) and unpaired t-tests.

## RESULTS

### Teeth statistics

For 736 possible teeth locations in all 23 cases, a total of 611 teeth were present (83%): 276 incisors (and canine), 176 pre-molars, and 159 molars. Figure 2 demonstrates d_mean_ distributions and missingness for individual teeth. The 3^rd^ molars were nearly always absent, with a quarter of the 2^nd^ molars missing. In general, the average d_mean_ was higher for all mandibular teeth compared to maxillary teeth, and d_mean_ was consistently higher for all molars compared to the incisors.

**Figure 2.**
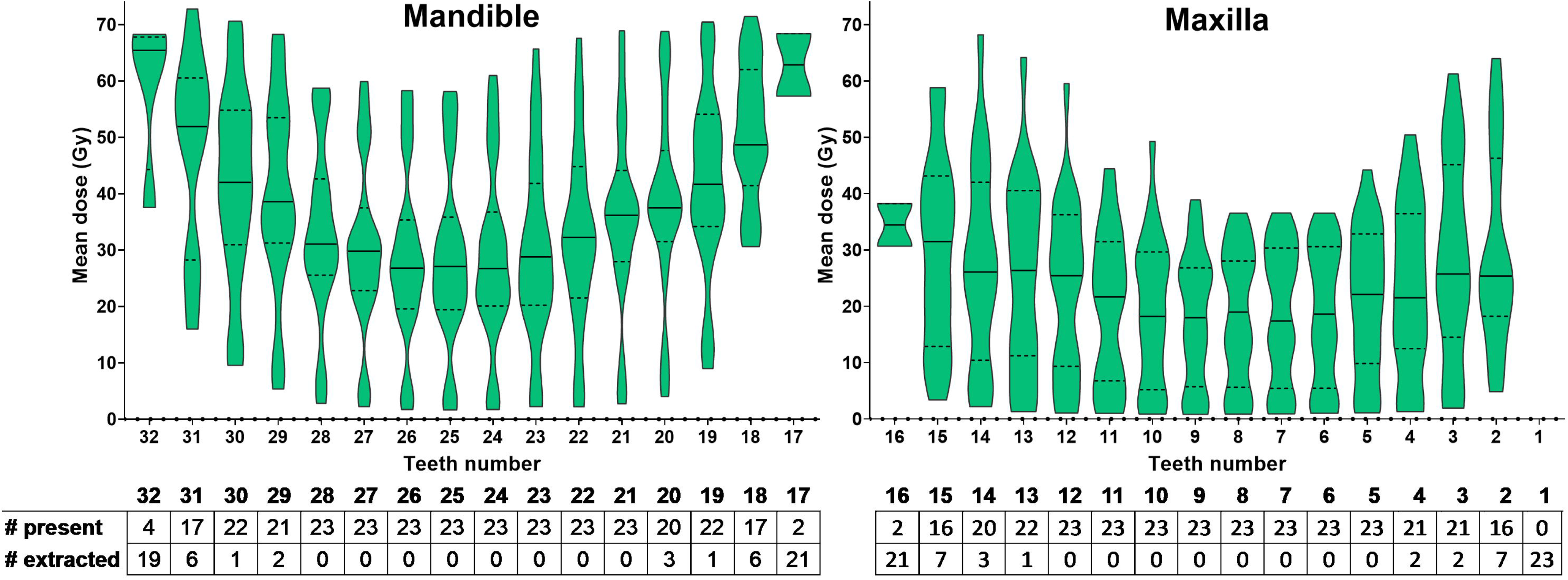
Violin plots of average tooth-based mean dose (d_mean_) for the mandible and maxilla in all 23 patients. The table below shows the number of teeth present in the cohort per tooth number. Violin plots are truncated at the minimum and maximum values.

### Manual tooth versus RADMAP jaw segment analysis

The mean geometric overlap between tooth roots and corresponding jaw segments was 77% [95% CI, 77–79%] without manual adjustment, rising to 90% [89–91%] after adjustment. By design, the alveolar jaw segments were larger (volume without/with adjustment: 1.16 cm^2^ [1.13-1.18] / 1.15 cm^2^ [1.12-1.18]) than manually delineated tooth roots (0.30 cm^2^ [0.29-0.31]), as they are intrinsically differently defined.

For *fully auto-segmented* alveolar jaw segments (Figure 3), there was a strong correlation between the d_mean_ to this jaw segment and the *manually* segmented teeth root (R^2^=0.98, p<0.0001). The Bland-Altman analyses showed a systematic d_mean_ bias of −0.7 Gy (SD: 2.3 Gy), indicating that the automated assessments of doses to the fully auto-segmented jaw segments tended to be higher than those of the manually segmented roots. The 95%-LOA ranged from –5.3 to 3.9 Gy. An opposite systematic bias was seen for teeth of the maxilla, as there was a positive systematic bias of 1.3 Gy (SD: 2.9 Gy), and 95%-LOA from −4.4 to 7.0 Gy.

**Figure 3.**
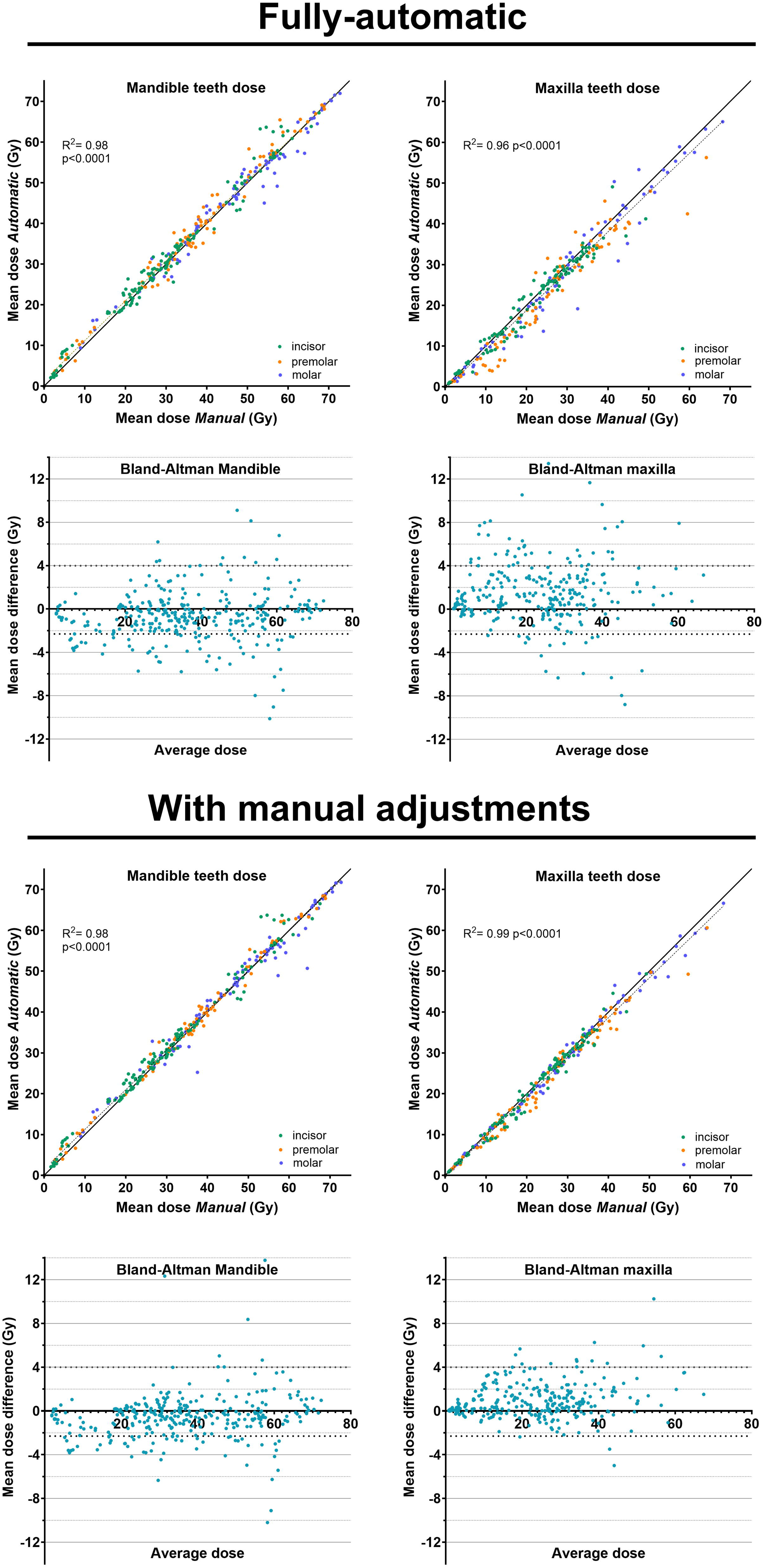
Radiation dose comparison plots between the fully-automatic teeth segments or with manual adjustments. The automatic versus manual d_mean_ for mandibular and maxillary teeth are demonstrated with color coding by incisor (green), premolars (orange), or molar (blue) teeth classification. Each comparative plot is accompanied by a corresponding Bland-Alman plot below.

For *auto-segmented* alveolar jaw segments *with adjustments* (Figure 3), there was again a strong correlation between the d_mean_ to this jaw segment (average of observers) and the *manually* segmented teeth root (R^2^=0.98, p<0.0001). The Bland-Altman analyses showed a slightly lower systematic bias of −0.6 Gy (SD: 2.2 Gy), as well as slightly narrower 95%-LOA: −4.8 to 3.7 Gy. The improvement of the manual adjustments with the tool compared to fully automatic was more pronounced for the maxilla, with a better agreement between the *manual* teeth and *auto-segmented* teeth region. The systematic mean teeth dose bias was of 0.8 Gy (SD: 1.6 Gy) with 95%-LOA from −2.3 to 4.0 Gy.

### Interobserver agreement

A total of 704 jaw segments were compared among all 6 observers (736 across 5 observers). The bland-Altman 95% LOA ranged from –2.88 to 2.88 Gy for the alveolar mandible segments and from –1.49 to 1.49 Gy for the maxilla, indicating that 95% of observer-derived dose values were within 2.88 Gy of the consensus d_mean_ (Supplement Figure S4. Bland-Altman plots). This interobserver analysis reflects comparisons between corresponding jaw segments, rather than between auto-segmented jaws and smaller tooth root areas. For the total jaw segments, results were comparable: the LOA was slightly narrower for the mandible (95%-LOA: −1.53 to 1.53 Gy) than the maxilla (95%-LOA: −1.52 to 1.52Gy). The mean systemic bias was negligible (≈0 Gy; <10_⁻_¹D), indicating no systematic over- or under-estimation among observers.

### ORN analyses

Of the 23 HNC patients, 11 (48%) developed ORN after RT. In total, 21 teeth regions were affected (6% of total teeth regions), and all were located in the mandible.

Figure 4 (top panel) shows that the d_mean_ to the entire mandible provided the least differentiation between patients who developed ORN and those who did not (difference in mean (DIM): 6.2 ± 4.3 Gy; p=0.17). Differentiation improved when using the manually segmented tooth roots (DIM: 12.9 ± 4.1 Gy; p = 0.0019), which was quite comparable for the alveolar jaw segments (DIM: 12.8 ± 3.9 Gy; p = 0.0012). The tooth-based total jaw segments also showed a significant distinction between ORN and non-ORN cases (DIM: 12.4 ± 3.7 Gy; p = 0.0008).

**Figure 4.**
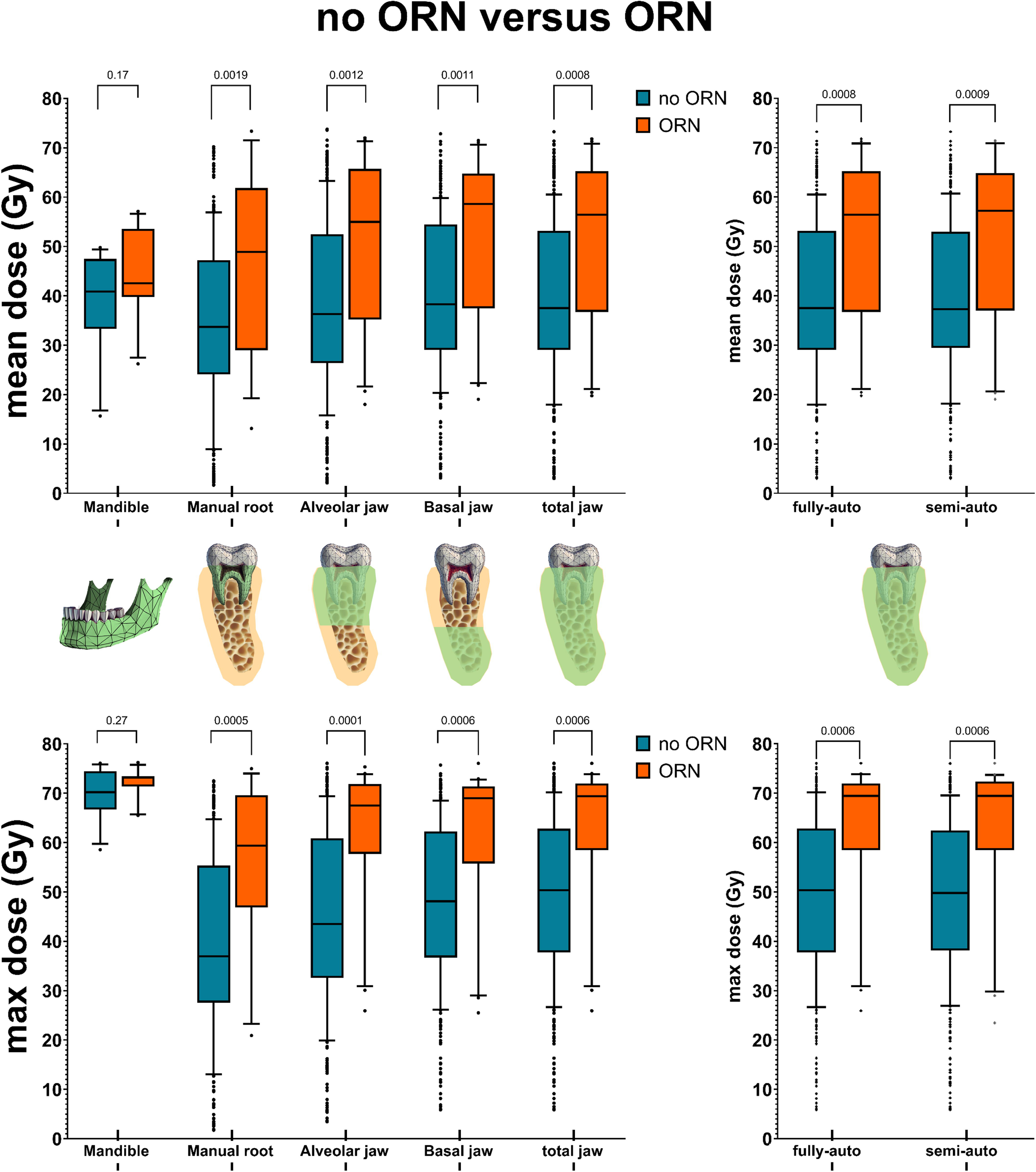
ORN distinction based on pre-treatment d_mean_ (top) and d_max_ (bottom) for the full mandible, manually segmented teeth root, and jaw segments (alveolar, basal and total). *Note: the sample size for p-value calculations are as follows: jaw subregions (n=368 teeth regions), roots (n=309 teeth regions), full mandible (n=23 full mandibles)*.

Figure 4 (bottom panel) shows that d_max_ generally yielded slightly better differentiation between ORN and non-ORN regions than d_mean_. The greatest distinction was observed for the alveolar jaw segments using d_max_ (DIM: 16.2 ± 4.1 Gy; p = 0.0001), while the total jaw segments also showed significant differences (DIM: 12.9 ± 3.7 Gy; p = 0.0006).

The fully automated and semi-automated jaw segmentation methods yielded similar differentiation between ORN and non-ORN regions for both the d_mean_ and d_max_ (Figure 4, right panels).

## DISCUSSION

This study introduces the RADMAP tool, designed to enhance multidisciplinary oral health care by integrating (semi-)automated, patient-specific dosimetric data—both quantitative and visual (i.e., dose maps)—into clinical decision-making. RADMAP divides the jaw into segments corresponding to individual tooth regions, making it independent of the number or presence of teeth at the start of RT, an important consideration since orodental sequelae often occur in edentulous jaw regions. Strong dose correlations were observed between RADMAP-defined tooth-specific jaw segments and manually segmented tooth roots for both fully and semi-automated segmentations. High interobserver agreement among six multidisciplinary users further demonstrated the tool’s reproducibility. Finally, radiation dose parameters from tooth-based jaw segments suggested superior differentiation between regions that later developed ORN and those that did not, compared with parameters derived from the entire jaw (current standard practice^6^). No difference in ORN differentiation was observed between the fully and semi-automated segmentations, suggesting that the tool may be used for this purpose without manual adjustments. By determining the radiation dose assessment per tooth-based jaw segment, RADMAP opens doors to improved orodental dose-toxicity modeling in patients receiving RT to the head and neck region.

Dental radiation dose mapping in head and neck cancer (HNC) radiotherapy has been explored previously. Polce et al. generated dental radiation-dose maps across 18 representative HNC-RT scenarios (i.e., glottic cancer, oropharynx, postoperative HNC), creating mean-dose tables for each tooth and each jaw third of the mandible and maxilla (i.e., lateral left, middle, and lateral right third).^24^ Their atlas approach expanded on earlier studies on dental dose distributions across HNC primary sites^25,26^ and modality comparisons (3DRT versus IMRT)^27^, but was limited by the assumption of uniform dose distributions across tumor sites and by lack of individualized segmentation. In contrast, RADMAP addresses this clinically unmet need by providing an RT planning-agnostic tool that intakes a patient-specific radiation dose distribution to (semi)automatically generate patient- and dentition-specific radiation dose maps. In addition to tabulated dose parameters, RADMAP produces a visual radiation odontogram that can be readily shared with dental practitioners to support coordinated pre- and post-RT oral care.

Manual delineation time of all 32 individual teeth and/or jaw segments remains a major barrier to implementing orodental dose reporting in clinical practice. In our study, full contouring time ranges from 35 to 120 minutes, depending on user experience. In contrast, minor manual adjustments (5–10 minutes) to the automated segmentation substantially improved geometric accuracy, increasing root inclusion from 77% to 90%. Despite intrinsic differences, manually defined tooth roots and alveolar jaw segments showed high correlation in radiation dose metrics, supporting the tool’s effectiveness for dose estimation in tooth-based jaw regions. Nevertheless, dose calculations were already highly consistent without adjustment and showed only slight improvement after refinement, underscoring the robustness and potential clinical utility of the automated approach. Moreover, RADMAP’s ray-based segmentations are independent of tooth presence, allowing accurate dose estimation even in edentulous regions. While patients in this study were selected based on having most teeth present, posterior molars were frequently absent (Figure 2). The tool nevertheless extracted dose parameters for these regions automatically. This is clinically relevant, as posterior jaw areas typically receive the highest radiation doses and are at greater risk of RT-related orodental sequelae.^7,25^

To our knowledge, this is the first study to evaluate ORN risk using spatially refined, tooth-based jaw segments and subregions (tooth root, alveolar, and basal bone), demonstrating better discrimination than whole-mandible dosimetry.^7^ Albeit the small patient numbers, our findings suggest that d_max_ may have a greater predictive capacity than d_mean_ for differentiating ORN from non-ORN sites. Additionally, ORN differentiation based on tooth-based jaw segment dosimetry was highly comparable between fully and semi-automated segmentations, supporting the reliability of the automated approach. Larger studies are needed to further investigate DVH associations with ORN development. Additionally, our tool enables the calculation of DVH parameters to teeth-based segments of the maxilla, where ORN may still occur, yet has been notoriously under-investigated in the literature.^28,29^

While mandible auto-segmentation is readily available in most HNC radiation oncology practices, maxilla segmentation is less common; while more common for non-RT cone-beam CT images.^30^ The open-source tool TotalSegmentator offers a solution for centers without dedicated autosegmentation software.^31^ Notably, RADMAP can also operate using mandible segmentation alone. To facilitate the usability of all features of our tool, we have also developed a DLC model for auto-segmentation of individual teeth, mandible, maxilla, and jaw substructures according to the new ORN ClinRad classification system^4,20^ which can be leveraged to generate orodental RT structure files (results not shown)^14^. As the RADMAP tool requires CT images and RT dose files as input, its clinical implementation will likely remain within radiation oncology. However, validation studies are needed to assess its usability among generalist radiation oncologists. The tool’s output is expected to be clinically relevant across disciplines—particularly for dental professionals to ascertain more precise radiation damage administered to the teeth regions, and adjust the dental follow-up accordingly — and can be readily shared through standard communication channels, for example as an image attachment in interdisciplinary correspondence.

## CONCLUSION

RADMAP is a (semi-)automated tool that generates patient-specific orodental dose maps by segmenting teeth-based regions of the mandible and maxilla directly from radiotherapy dose distributions. It provides clinically relevant radiation-dose reporting in both tabular and visual (odontogram) formats. Dosimetric analyses suggested potential superior tooth-level differentiation of ORN risk compared with whole-jaw dose assessment, even without manual adjustment. The RADMAP tool is patient-specific, agnostic to treatment planning software, and independent of missing teeth. Tooth-level dose assessment with RADMAP improves communication between radiation oncology and oral and dental health care teams and provides a foundation for future modeling of radiation-related orodental toxicities.

## Data Availability

All data produced are available online at

https://figshare.com/s/816bca5756ad22047bfe

## Data Sharing Statement

In accordance with the *Final NIH Policy for Data Management and Sharing* NOT-OD-21-013, data that support the findings of this study are openly available in an NIH-supported generalist scientific data repository (figshare) at 10.6084/m9.figshare.30465716 no later than the time of an associated publication; while public data is embargoed pending peer review, referees may access this data via private link at https://figshare.com/s/816bca5756ad22047bfe during the peer review process.

## CRediT statement

Conceptualization: LVD, LHV, ACM, CDF, RAAW, MSC, NAM, MJHW. Methodology: LVD, LHV, ACM, CDF, MSK, MSC, NAM, HCW, RH, AH. Software: LVD. Validation: LVD, LHV, ACM, EEW, RAAW, LEJ. Formal analysis: LVD, LHV, ACM. Investigation: LVD, LHV, ACM, EEW, RAAW, LEJ. Resources: LHV, ACM, CDF, SYL. Data curation: LHV, ACM, RDJ, EW, NAM, HCW, RH. Writing – original draft: LVD, LHV, ACM, CDF, MSK. Writing – review and editing: LVD, LHV, ACM, CDF, EEW, RAAW, LEJ, MSK, SYL, RDJ, MSC, NAM, HCW, RH, AH, MJHW. Visualization: LVD, LHV, ACM. Funding acquisition: ACM, CDF.

