## Supplementary data for "Radiation-specific Automated Dosimetric dental, Mandible, and maxilla Annotation for Predicting Periodontal Problems (RADMAP): A semi-automated tool for dosimetric oral risk communication and osteoradionecrosis assessment"

### Supplementary material

**Supplement S1. CT DICOM header information**

| Manufacturer | ManufacturerModelName | SliceThickness | PixelSpacing | Exposure |
|---|---|---|---|---|
| GE MEDICAL SYSTEMS | Discovery ST | 3,75 | 0,98 | 27 |
| Philips | Brilliance 64 | 2,5 | 0,98 | 175 |
| Philips | Mx8000 IDT 16 | 3 | 0,98 | 300 |
| SIEMENS | SOMATOM Definition Edge | 3 | 1,04 | 262 |
| Philips | Mx8000 IDT | 3 | 0,98 | 350 |
| Philips | Brilliance 64 | 2,5 | 0,98 | 250 |
| Philips | Brilliance Big Bore | 3 | 1,17 | 410 |
| Philips | Mx8000 IDT 16 | 3 | 0,98 | 300 |
| Philips | Brilliance 64 | 2,5 | 0,98 | 175 |
| GE MEDICAL SYSTEMS | LightSpeed RT | 2,5 | 0,98 | 221 |
| SIEMENS | SOMATOM Definition Edge | 2 | 0,98 | 500 |
| GE MEDICAL SYSTEMS | LightSpeed RT16 | 2,5 | 0,98 | 30 |
| SIEMENS | SOMATOM Definition Edge | 2 | 0,98 | 500 |
| SIEMENS | SOMATOM Definition Edge | 2 | 0,98 | 500 |
| Philips | Brilliance Big Bore | 3 | 1,04 | 420 |
| Philips | Mx8000 IDT 16 | 3 | 0,98 | 300 |
| Philips | Brilliance Big Bore | 3 | 1,36 | 376 |
| Philips | Brilliance Big Bore | 3 | 1,04 | 450 |
| Philips | Brilliance Big Bore | 3 | 1,17 | 425 |
| Philips | Brilliance Big Bore | 3 | 1,37 | 376 |
| Philips | Mx8000 IDT 16 | 3 | 0,98 | 300 |
| Philips | Brilliance 64 | 2,5 | 0,98 | 175 |
| Philips | Brilliance 64 | 1 | 0,98 | 175 |

**Supplement S2. Delphi-study results**

Visualized information and functionality of the RADMAP tool was influenced by the results of an international expert Delphi study (n=69) that developed consensus-based guidelines for key data elements of ORN.[16] During rounds 1 and 2 of the study, experts were asked what information was deemed useful to present in a tool that communicated radiation dose distributions to teeth and teeth-bearing regions of the jaw. Nearly 90% (51/57) found a ‘heat map’ of radiation dose over an odontogram clinically useful and 81% found it somewhat or very easy to interpret (46/57). There was greater preference (59%, 33/56) to visualize the odontogram as an open mouth structure rather than in chart form, and over 96% (53/55) desired the inclusion of dose data to tooth-bearing (i.e., segmented jaw regions) where teeth were missing in the RADMAP output for each patient. Out of 5 different tooth-based dosimetric parameters presented (i.e., max point dose [$d_{max}$], mean dose [$d_{mean}$], dose going to 0.03cc, dose going to 95% of the tooth [D95], or D50), $d_{max}$ and $d_{mean}$ were ranked the highest in preference.

**Supplement Figure S3**. RADMAP tool – in and output

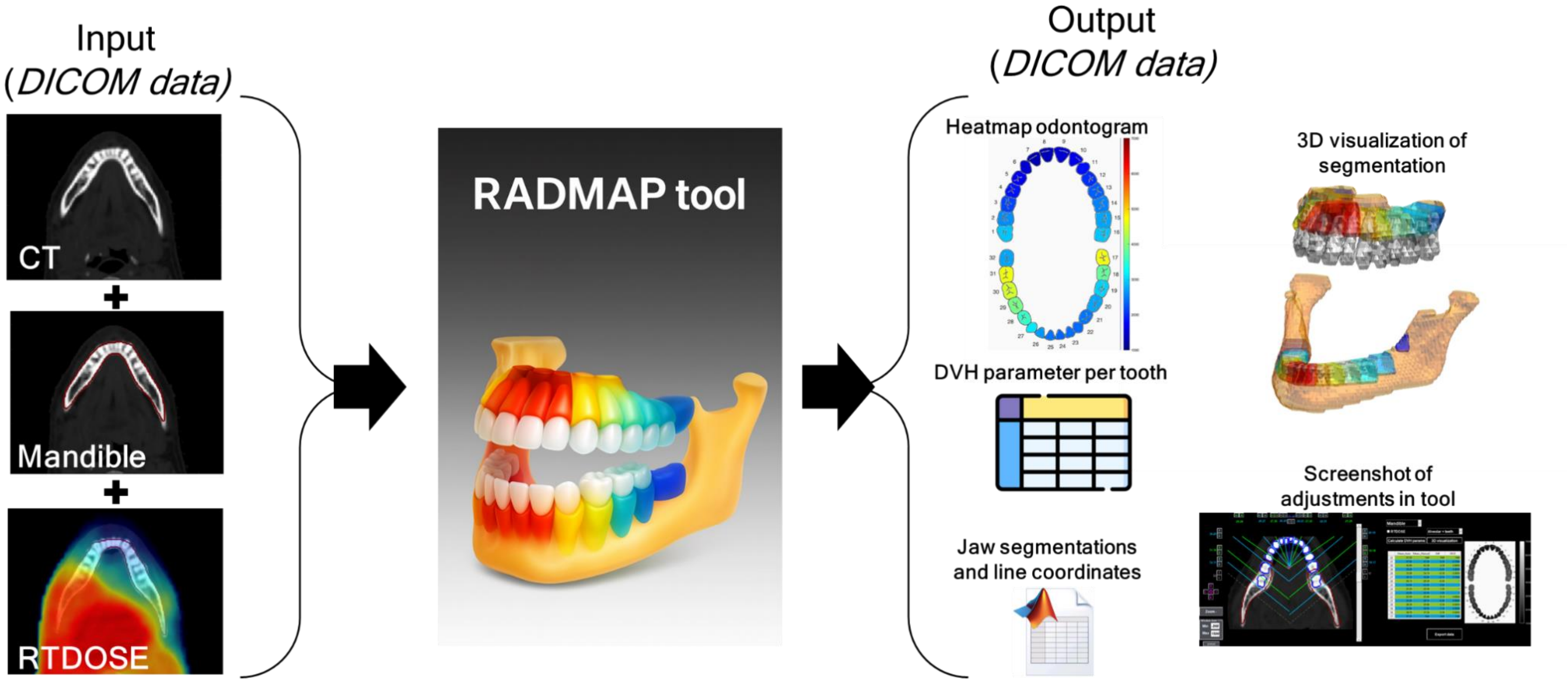

**Supplement Figure S4.** Visualization of entire mandible, manually segmented tooth roots, and fully auto-delineated jaw, alveolar, and basal bone segments

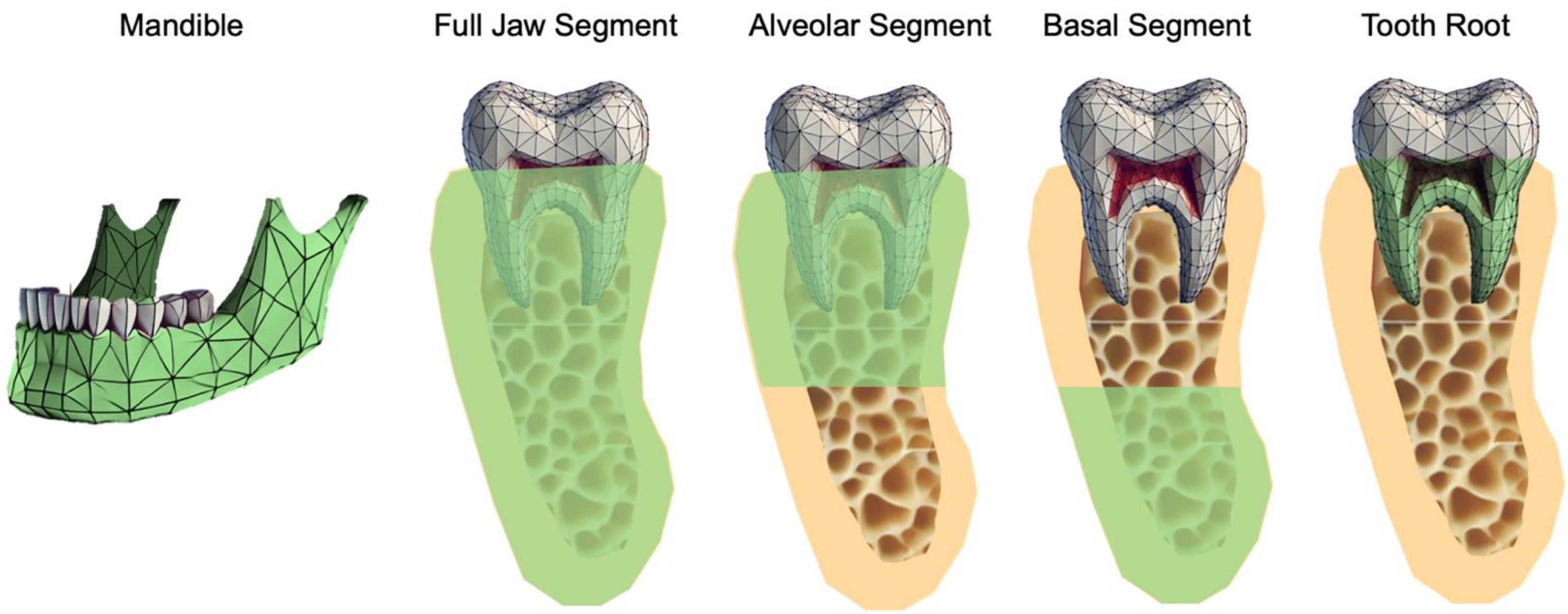

**Supplement Figure S5.** Bland-Altman - Interobserver analysis

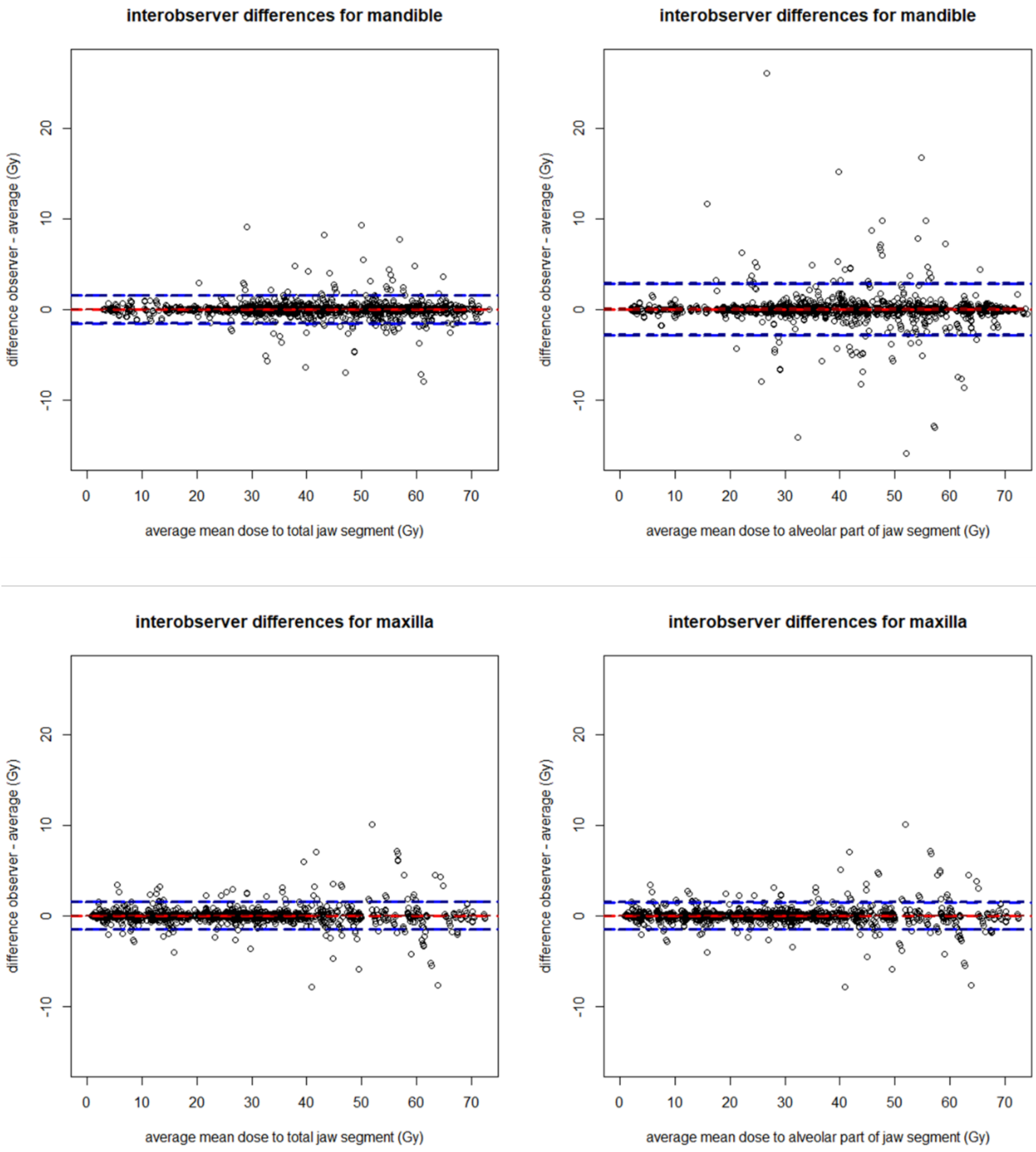
